# A genotype-first approach identifies high incidence of *NF1* pathogenic variants with distinct disease associations

**DOI:** 10.1101/2023.08.08.23293676

**Authors:** Anton Safonov, Tomoki T Nomakuchi, Elizabeth Chao, Carrie Horton, Jill S Dolinsky, Amal Yussuf, Marcy Richardson, Virginia Speare, Shuwei Li, Zoe C Bogus, Maria Bonanni, Anna Raper, Staci Kallish, Marylyn D Ritchie, Regeneron Genetics Center, Penn Medicine BioBank, Katherine L Nathanson, Theodore G Drivas

## Abstract

Loss of function variants in the *NF1* gene cause neurofibromatosis type 1 (NF1), a genetic disorder characterized by complete penetrance, prevalence of 1 in 3,000, characteristic physical exam findings, and a substantially increased risk for malignancy. However, our understanding of the disorder is entirely based on patients ascertained through phenotype-first approaches. Leveraging a genotype-first approach in two large patient cohorts, we demonstrate unexpectedly high prevalence (1 in 450-750) of *NF1* pathogenic variants. Half were identified in individuals lacking clinical features of NF1, with many appearing to have post-zygotic mosaicism for the identified variant. Incidentally discovered variants were not associated with classic NF1 features but were associated with an increased incidence of malignancy compared to a control population. Our findings suggest that *NF1* pathogenic variants are substantially more common than previously thought, often characterized by somatic mosaicism and reduced penetrance, and are important contributors to cancer risk in the general population.

## Introduction

The human disease gene *NF1* encodes the protein neurofibromin, a tumor suppressor and negative regulator of the RAS/MAPK pathway.^1–4^ Heterozygous loss of function variants within the *NF1* gene lead to the autosomal dominant disorder neurofibromatosis type 1 (NF1), with an estimated prevalence of 1 in 3,000.^1–4^ NF1 is often cited as a classic example of a pleiotropic genetic disorder with variable expressivity and complete penetrance; that is, every individual with a germline pathogenic *NF1* variant is expected to meet NF1 diagnostic criteria, although their specific presentation and degree of organ involvement may be variable.^2–5^ The classic features as defined by the NIH clinical diagnostic criteria include: cutaneous café-au-lait macules (CALMs), neurofibromas, axillary/inguinal freckling, iris hamartoma, and certain skeletal anomalies.^6^ Affected individuals are also at increased risk for hypertension, cardiovascular anomalies, and certain malignancies, making early diagnosis and screening a critical aspect of care.^4^

The malignancies classically seen in NF1 patients are of neural crest derivation, however patients with NF1 have also been observed to have increased risk for a broad range of different malignancies, often with distinctive disease behavior and prognosis.^7, 8^ For example, breast cancers in female patients with NF1 are thought to be characterized by an earlier age of onset, increased mortality, and unfavorable prognostic factors, such as estrogen/progesterone receptor negativity and *HER2* amplification.^8–10^ With the advent of widespread tumor sequencing in cancer patients without NF1, it has become clear that somatic *NF1* driver mutations are also common in certain tumors, including melanoma, glioblastoma, and breast and ovarian cancer.^11^

Somatic mosaic *NF1* variants have also rarely been reported in clonal hematopoiesis (CH),^12, 13^ an age-and oncologic treatment-related phenomenon characterized by the presence of clonal, genetically distinct subpopulations of the hematopoietic lineage within a single patient,^12, 14^ and in the condition known as segmental neurofibromatosis, a subtype of NF1 with a reported incidence of roughly 1 in 75,000.^15^ Segmental NF1 results from a somatic post-zygotic mutation early during embryogenesis, with the resultant mutant cell lineage going on to populate limited areas of the body which manifest as foci of affected tissue displaying classic NF1-associated features, and with the causal *NF1* variant detectable only in affected cells. On the other hand, CH results from somatic mutations occurring late in life and only in the blood, and is therefore not expected to be associated with classic NF1 presentations.^16, 17^

With the advent of broadly applied next-generation sequencing technologies and multi-gene panel testing (MGPT), incidental, mosaic, and otherwise unexpected genetic findings are becoming more common;^18–20^ the identification of an incidental germline pathogenic *NF1* variant in a patient with breast cancer without a clinical NF1 diagnosis would be one such example. Similarly, as the scientific community develops large-scale population-level biobanks,^21, 22^ we will identify previously undiagnosed individuals with pathogenic variants in disease genes.^20, 23^ Medical management of patients with incidentally discovered genetic variants can prove problematic as we currently lack clear clinical guidelines for their appropriate care and counseling, and the clinical relevance of an incidentally discovered variant in the absence of a congruent clinical phenotype is unknown.

To investigate the prevalence of *NF1* pathogenic variants on a population-scale, we evaluated two cohorts of individuals from independent datasets: the population-level Penn Medicine Biobank (PMBB, n = 43,731) and a database of patients clinically sequenced for cancer risk evaluation by Ambry Genetics (n = 118,768). We identified an unexpectedly high prevalence (1 in 450-750) of pathogenic variants in the *NF1* gene, more than four times the rate expected given the reported prevalence of NF1. Half of these individuals lacked any evidence of syndromic NF1, and many, but not all, appeared to be mosaic for the identified *NF1* variant. The discovery of an incidental *NF1* pathogenic variant did not correlate with the presence of classic symptoms of NF1 but was associated with a greater incidence of certain malignancies compared to a matched control population. Our findings suggest that *NF1* pathogenic variants are substantially more common than previously thought, often characterized by somatic mosaicism and reduced penetrance, and are important contributors to cancer risk in the general population.

## Results

### Patients with incidentally discovered *NF1* pathogenic variants have no evidence of NF1

The University of Pennsylvania Department of Medicine, Division of Translational Medicine and Human Genetics, has evaluated four patients, Cases 1-4 (**Table S1**), for NF1, all of whom had been incidentally diagnosed with an *NF1* pathogenic/likely pathogenic variant (*NF1* PVs). In all four cases comprehensive physical exam, medical history, and family history revealed no or very few features consistent with an NF1 diagnosis; none of these four cases met diagnostic criteria for NF1,^24^ contrary to the reported complete penetrance of the disorder, and despite genetic data strongly suggestive of heterozygous germline variation for the *NF1* variant in at least one individual.

### Frequency of *NF1* PVs in two large patient cohorts

To investigate the frequency and penetrance of *NF1* PVs in a more unbiased way, we utilized the Penn Medicine BioBank (PMBB), a large academic medical biobank with exome sequencing data on 43,731 individuals, all patients of the University of Pennsylvania Health System (UPHS).^25^ We identified 58 individuals heterozygous for any of 50 unique *NF1* PVs: 43 predicted loss of function (pLOF) variants, five missense variants, and two deletions involving the entire *NF1* gene (**Figure 1A, Table S2**). This prevalence of 1 in 752 (0.13%) is four-fold greater than the reported prevalence of 1 in 2,500 to 3,500 for NF1.^2–4^

**Figure 1.**
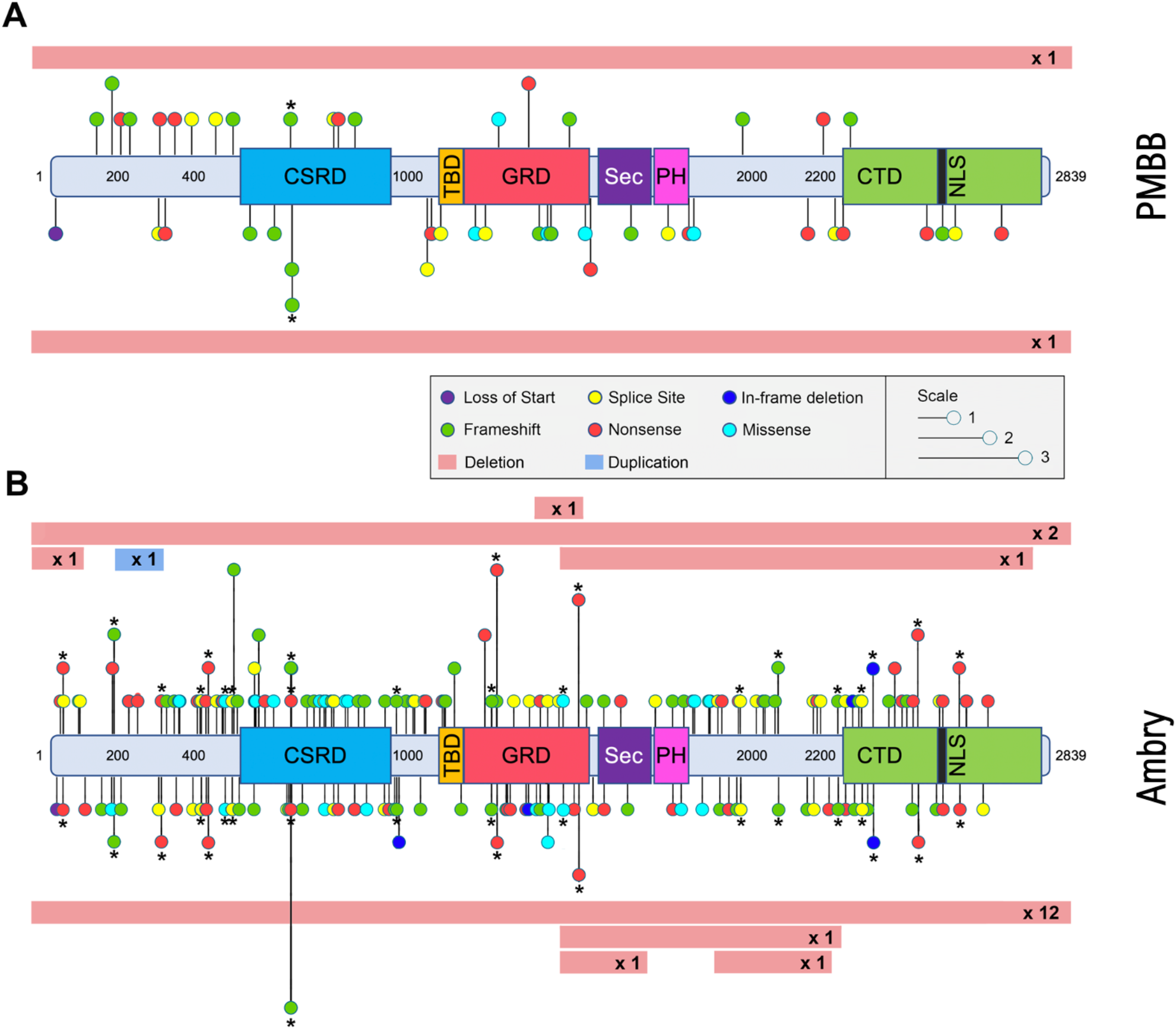
*NF1* variants identified in the present study. The *NF1* variants identified in (A) the PMBB dataset and (B) the Ambry dataset are displayed along a schematic of the NF1 protein. For both *NF1* variants identified in the Clinical-NF1 group are indicated along the top of the protein schematic, whereas those identified in the PV-Only group are indicated along the bottom. Variants labeled with an asterisk were identified in both Clinical-NF1 and PV-Only individuals within each dataset. Variants are color-coded by predicted protein effect, with the height of each line segment corresponds to the number of individuals in which the indicated variant was identified. Large deletions and duplications are indicated by red and blue bars, respectively. Amino acid position, based on NP_001035957, are indicated along the protein schematic. NF1 protein domains are indicated, as follows: CSRD, Cysteine-and-Serine-Rich Domain; TBD, Tubulin-Binding Domain; GRD, GAP-Related Domain; Sec, Sec14 Homologous Domain; PH, Pleckstrin Homologous Domain; CTD, C-terminal Domain; NLS, Nuclear Localization Signal.

We replicated our analysis in a cohort of 118,769 patients collected by Ambry Genetics, all of whom had undergone multi-gene panel testing (MGPT) for hereditary cancer predisposition with gene panels including the *NF1* gene from January 2014 through March 2018 (**Table S3**). We identified 281 individuals heterozygous for any of 219 unique *NF1* PVs: 170 pLOF variants, 24 missense variants, 10 exonic deletions/duplications, three single amino acid deletions, and 12 deletions involving the entire *NF1* gene (**Figure 1B, Table S4**). No patient had multiple *NF1* PVs identified. This prevalence was 1 in 432 individuals (0.24%).

### Half of *NF1* PV carriers do not have an NF1 diagnosis

We completed a chart review of all 58 patients with an *NF1* PV within the PMBB cohort, specifically looking for clinical evidence of NF1. Only 23 of the 58 *NF1* PV carriers (39.7%) had a known diagnosis of NF1. We thus divided the PMBB cohort into two groups: the Clinical-NF1 group (those with an *NF1* PV and a known NF1 diagnosis) and the PV-Only group (those with an *NF1* PV but without a known diagnosis of NF1). Only two of the 35 PV-Only individuals had any medical history consistent with NF1 (**Table S2**). Individual P39 was noted to have CALM on skin exam and had severe upper limb deformities and spina bifida; a diagnosis o f NF1 had not been made. Individual P38 was noted to have multiple nevi on skin exam; interestingly, this individual had also been diagnosed with bilateral retinoblastoma in childhood and had been found to harbor a pathogenic *RB1* variant on clinical genetic testing. No other patients in the PV-Only group had reported evidence of any other unusual dermatologic findings, axillary/inguinal freckling, or neurofibromas. Individual P41 in the Clinical-NF1 group had been evaluated in our clinic and was given a clinical diagnosis of NF1 based on widespread CALM and innumerable neurofibromas, with all dermatomes apparently affected. However, clinical genetic testing of the *NF1* gene in this patient returned negative, while research-based sequencing in PMBB revealed an *NF1* PV with a variant allele fraction of 0.10, suggesting post zygotic mosaicism for the variant. Lastly, chart review revealed that individual P32 was the same individual that had been referred to our clinic as Case 1, reported above, with no evidence of NF1 on our own detailed physical exam.

Dividing the Ambry cohort into the same Clinical-NF1 and PV-Only groups, we observed very similar results. Although we did not have the same depth of phenotypic information available to us in the Ambry cohort as in PMBB, using a combination of physical exam reports, family history, clinic notes, test requisition forms, and outreach to ordering providers (**Figure S1**, **Supplemental Results**), we were able to classify 152 of the 281 *NF1* PV carriers (54%) as Clinical-NF1, whereas 129 patients (46%), lacking any evidence of a known NF1 diagnosis, were classified as PV-Only (**Table S4**). Thus, 48.7% of *NF1* PV variant carriers across both cohorts, appeared to lack a diagnosis of NF1.

### Significant demographic differences but no differences in medical comorbidities between the Clinical-NF1 and PV-Only groups

Analysis of demographic and medical history differences between the Clinical-NF1 and PV-Only groups (**Table S5-6**) re vealed that the Clinical-NF1 group was significantly younger than the PV-Only group in both the PMBB and Ambry cohorts (PMBB, mean age of Clinical-NF1 patients 45.0 years, mean age of PV-Only pat ients 66.1 years, p=1.37e-7; Ambry, mean age of Clinical-NF1 patients 46.3 years, mean age of PV-Only patients 55.9 years, p=1.54e-08). In the PMBB cohort, patients in the Clinical-NF1 group were less likely to self-identify as White compared to the PV-Only group (p=9.42e-3) – this difference did not replicate in the Ambry dataset. In PMBB we did not identify any significant differences in any major medical comorbidities or anthropometrics between the two groups aside from a nominally significant increased rate of anxiety/depression in the Clinical-NF1 group (**Table S5**); adjusting models for sex and age, there were no significant differences in height (p=0.209), incidence of hypertension (p=0.85), history of malignancy (p=0.30), or any other common or NF1-associated diagnoses (**Table S5**).

### No difference in the types of *NF1* PVs between the Clinical-NF1 and PV-Only groups

Classifying *NF1* PVs by predicted protein effect, we observed a statistically significant enrichment of whole-gene deletions in the PV-Only group compared to the Clinical-NF1 group in the Ambry cohort (p=0.008, **Figure S2A-B, Table S6**); in total, 10 whole-gene deletions were found among the PV-Only group (representing 8.2% of all variants found in this group), whereas only two were found in the Clinical-NF1 group (representing 1.3% of all variants found in this group). We did not observe this phenomenon in PMBB (**Figure S2A-B, Table S5**), with only two whole-gene deletions identified, one in the PV-Only group and one in the Clinical-NF1 group. No other significant differences were seen in predicted *NF1* variant effect in either cohort. No differences were seen between the nature of the nucleotide change between the Clinical-NF1 and PV-Only groups in either PMBB or Ambry; in both cases C>T transitions were by far the most common (**Figure S2C-D**).

### Evidence for somatic mosaicism in the PV-Only groups

In PMBB, the mean variant allele fraction (VAF) of the *NF1* PVs identified in the Clinical-NF1 group was 0.47, consistent with heterozygous germline variation. On the other hand, the PMBB PV-Only group had a mean *NF1* PV VAF of 0.29 (**Figure 2A, Table S3**). This statistically significant difference (p=4.54e-06) suggesting the presence of somatic mosaicism in at least some individuals in the PV-Only group.

**Figure 2.**
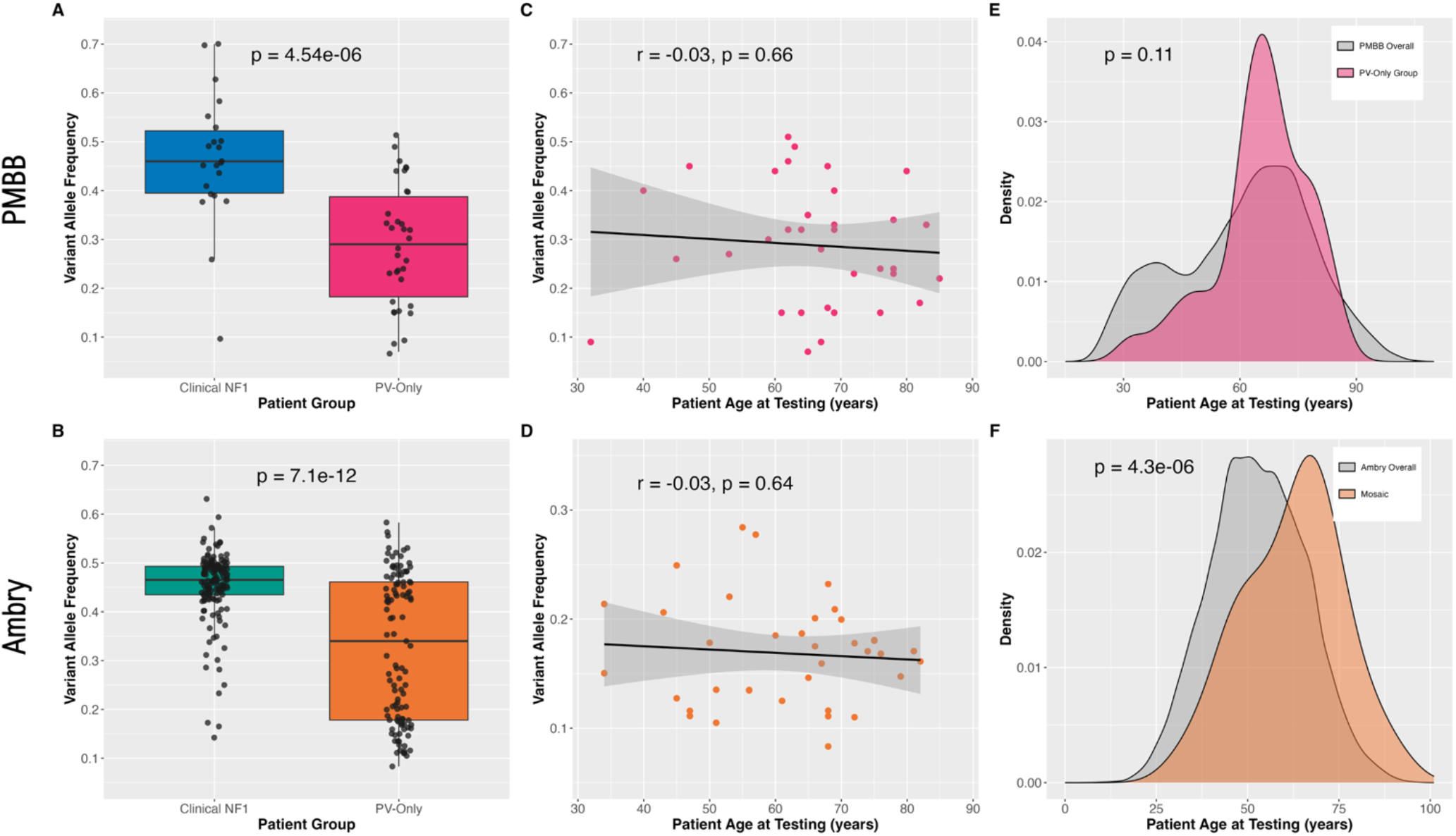
Characterization of somatic mosaicism of *NF1* PVs in the PMBB and Ambry datasets. (A) The variant allele fraction (VAF) for each *NF1* PV identified in individuals in the Clinical-NF1 group (left, blue) and PV-Only group (right, pink) are displayed for PMBB. Box plots illustrate the median, first and third quartiles, minimum, and maximum for each group. The difference in means between the two groups was statistically significant by linear regression (p=4.54e-06). (B) Comparison of *NF1* PV VAF, as in panel A, but for the Ambry data; individuals in the Clinical-NF1 group are shown on the left in green and individuals in the PV-Only group are shown on the right in orange. The difference in means between the two groups was statistically significant by linear regression (p=7.1e12). (C) Linear regression of patient age, in years (horizontal axis), against *NF1* PV VAF (vertical axis) for the PMBB PV-Only group. Each point represents a single individual. The regression line is shown, with grey shading illustrating the 95% confidence intervals. There is no significant correlation between the two variables (Pearson correlation coefficient of -0.03, p=0.66). (D) Linear regression of patient age against *NF1* PV VAF as in panel B, but for the 47 Ambry PV-Only patients with a mosaic *NF1* PV. There is no significant correlation between the two variables (Pearson correlation coefficient of -0.02, p=0.64). (E) The distribution of patient ages at time of genetic testing, in years (horizontal axis) for all 43,559 PMBB participants without an *NF1* PV (grey) and for the 35 individuals in the PMBB PV-Only group (pink). There is no significant difference in the distribution of patient ages between the two groups by the Wilcoxon rank sum test (p=0.11). (F) The distribution of patient ages at time of genetic testing for all 118,709 Ambry patients tested with MGPTs containing the *NF1* gene (grey) and the 46 individuals in the Ambry PV-Only group with confirmed mosaic *NF1* PVs (orange). The Ambry PV-Only group with mosaic *NF1* PVs is significantly older than the overall Ambry cohort by the Wilcoxon rank sum test (p=4.3e-06).

However, not all individuals in the PV-Only group had low VAF *NF1* variants; 17 of the 35 individuals (49%) had *NF1* PVs with VAF ≥ 0.3 (**Figure 2A, Table S2**), more suggestive of heterozygous germline variation. Additionally, two of the 23 Clinical-NF1 individuals (8.7%), including individual P41 discussed above, had an NF1 PV VAF < 0.3 (range: 0.10 – 0.26), suggestive of mosaicism despite their clinically affected status.

In the Ambry cohort, we found a mean *NF1* PV VAF of 0.45 in the Clinical-NF1 group, again consistent with heterozygous germline variation, whereas the PV-Only group was found to have a mean VAF of 0.35 (p=7.1e-12), suggestive of somatic mosaicism (**Figure 2B, Table S6**). Again, not all individuals in the PV-Only group were found to have a reduced VAF *NF1* PV; the VAF within the PV-Only group had a bimodal distribution (**Figure 2B**), suggesting the existence of two populations, with 58 (45%) individuals having VAF ≥ 0.3, 53 (41%) with VAF < 0.3, and 18 (14%) for whom VAF was unavailable (mostly due to technical limitations of copy number variant analysis). Of the 18 individuals for whom VAF was not available, eight were found to be likely mosaic for their *NF1* PV by Sanger sequencing. Of the 53 *NF1* PV-Only samples with VAF < 0.3, 39 (74%) also had evidence of mosaicism on confirmatory Sanger sequencing (data not shown). Thus, altogether 47 (39 with low VAF by NGS, confirmed by Sanger sequencing, eight without VAF from NGS but likely mosaic by Sanger sequencing) of the 129 individuals in the Ambry PV-Only group (36%) were likely mosaic for their *NF1* PV and are indicated as “mosaic” in **Table S5**. Additionally, within the Ambry Clinical-NF1 group, seven individuals (4.6%) were found to have an *NF1* PV VAF < 0.30 (range: 0.14 – 0.29, further clinical and molecular characteristics regarding these cases is included in **Table S7**), but only one of these seven also had evidence of reduced allelic fraction based on Sanger sequencing.

Comparing the predicted protein effect of *NF1* PVs between the germline and mosaic individuals in both PMBB and Ambry (**Figure S2B,C**), we saw that the enrichment of *NF1* whole gene deletions that we had observed in the Ambry PV-Only group was accentuated in the mosaic *NF1* PV group, with 12.8% of all variants in the mosaic *NF1* PV group found to be whole gene deletions, compared to only 2.5% in the heterozygous *NF1* PV group (p=0.0015). No other significant differences were seen.

### Somatic mosaicism of *NF1* PVs and patient age

Recent studies have rarely identified somatic mosaic *NF1* variants in patients with CH.^12, 13^ We investigated the possibility of CH as a driver of *NF1* PV mosaicism in the PMBB cohort, plotting patient age against *NF1* PV VAF for all individuals within the PV-Only group (**Figure 2C)**. We found no correlation between patient age and *NF1* PV VAF (r=-0.03, p=0.66), contrary to what would be expected in CH. Replicating this analysis in the Ambry cohort, we again found no correlation between patient age and *NF1* PV VAF (**Figure 2D**, r=-0.03, p=0.64), again arguing against CH as a mechanism of mosaicism. Since the incidence of CH increases sharply with patient age,^12, 14^ we asked whether patients in the PV-Only groups might be significantly older than the overall study populations from which they were drawn. In PMBB, no difference was seen between the ages of the PV-Only individuals and the ages of the 43,559 individuals in the overall PMBB population (**Figure 2E**, p=0.11 by Wilcoxon rank sum test), again contrary to what would be expected in CH. On the other hand, within the Ambry cohort, the PV-Only individuals with mosaic *NF1* PVs were significantly older than the overall Ambry population of 118,709 patients (**Figure 2F**, Wilcoxon Rank Sum test p=4.3e-06). Together these data suggest that an age-related process such as CH likely contributes to but is not entirely responsible for the mosaicism that we observed.

### PheWAS analysis in PMBB for NF1-associated phenotypes

Leveraging the deep phenotypic data available for PMBB participants, we completed a Phenome-Wide Association Study (PheWAS) across 9,030 ICD-10 code-based phenotypes to discover, in an unbiased way, patient phenotypes significantly associated with the presence of an *NF1* PV and identified 53 significant associations (p<5.3e-6, **Figure 3A, Table S8**). The most statistically significant associations were for the ICD-10 codes Q85.00 (Neurofibromatosis, unspecified) and Q85.01 (Neurofibromatosis, type 1). The remaining 51 significant associations, all known features of syndromic NF1, were for ICD-10 codes broadly characterized by benign/malignant neoplasms, leukemia, stress fracture, and scoliosis (**Figure 3A, Table S8)**. A number of other phenotypes that have previously been suspected of being associated with NF1 came close to reaching significance, including interstitial emphysema (p=5.95e-5) and functional diarrhea (p=6.33e-5)^26–28^.

**Figure 3.**
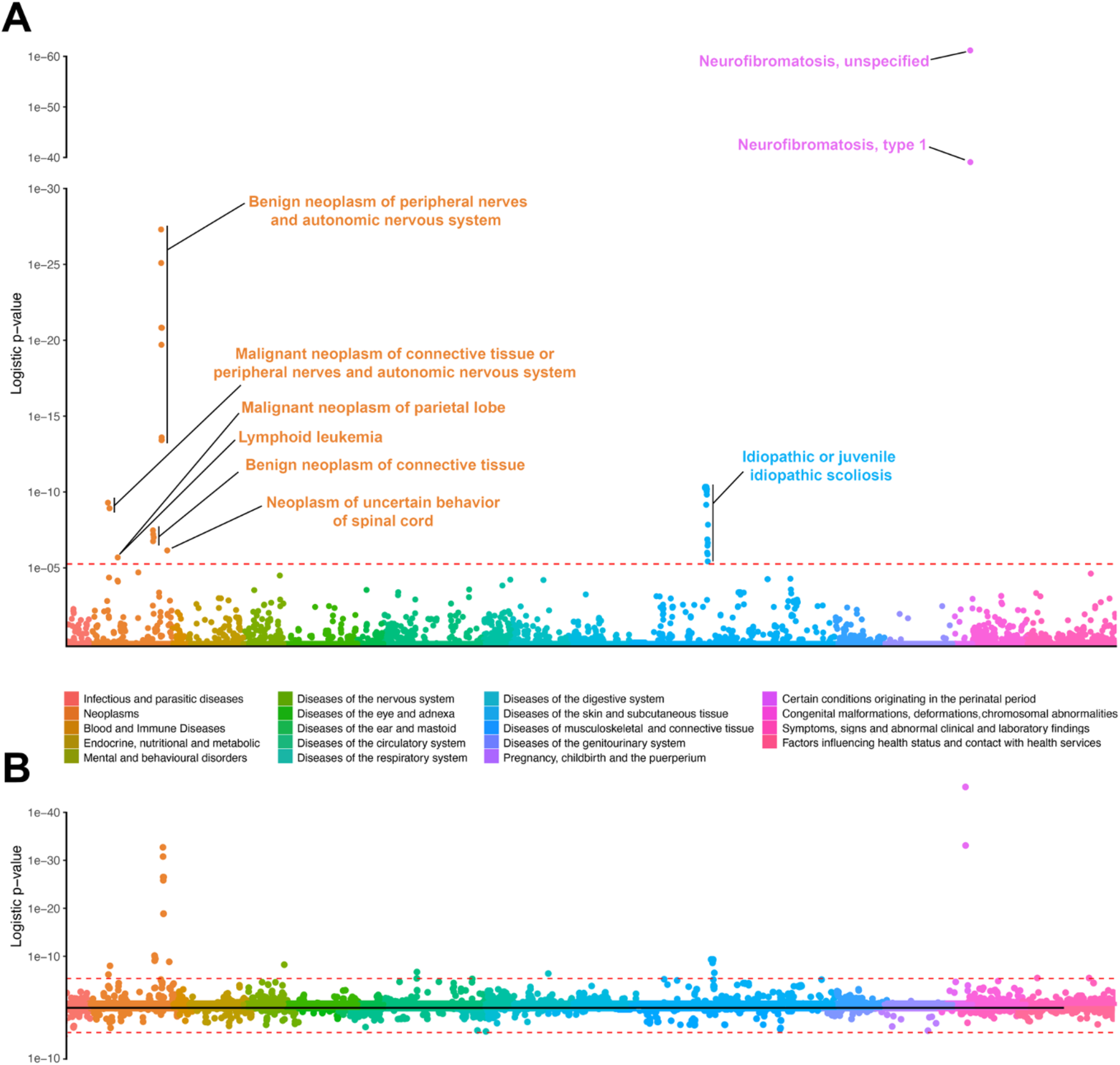
PheWAS Results for *NF1* PV carriers in the PMBB cohort. Phenome-wide association studies (PheWAS) were performed to identify ICD-10 code phenotypes significantly enriched within *NF1* PV carriers in PMBB. In panels A and B, individual ICD-10 phenotypes are indicated along the horizontal axis with each point (colored by phenotype group) representing a single ICD-10 code. The height of each point along the vertical axis corresponds to the strength of association for that phenotype with *NF1* PV carrier status, with the p-value of association indicated along the vertical axis. The Bonferroni-corrected p-value significance threshold of 5.5e-6 (correcting for testing across 9,436 individual ICD-10 codes) is indicated by red horizontal dashed lines. (A) PheWAS results for all 58 *NF1* PV carriers in PMBB are displayed. Phenotype associations surpassing the Bonferroni-corrected significance threshold are labelled (ICD-10 codes with similar descriptions are labeled as groups; more detailed results can be found in **Table S8**). (B) The top portion of the Miami plot illustrates results of PheWAS analysis excluding the 35 *NF1* PV carriers in the PV-Only group; the bottom portion of the plot illustrates results of repeat PheWAS analysis excluding the 23 *NF1* PV carriers in the Clinical-NF1 group. More detailed results can be found in **Table S9-10**.

We repeated the PheWAS but excluded either the 35 PV-Only individuals (**Figure 3B**, top panel, **Table S9**) or the 23 Clinical-NF1 individuals from analysis (**Figure 3B**, bottom panel, **Table S10**). PheWAS results considering only the Clinical-NF1 individuals identified 43 statistically significant phenotypic associations, of which, 39 (89%) had also been identified in our initial analysis of all 58 *NF1* PV carriers. On the other hand, PheWAS results considering only the PV-Only group, identified no significant disease associations. With the caveat that this sub-analysis is relatively underpowered, these results suggests that the presence of an incidentally discovered *NF1* PV in blood confers little risk for phenotypes classically associated with syndromic NF1.

### Increased incidence of malignancy in both the PV-Only and Clinical-NF1 groups

Within the Ambry cohort, 20 of the 281 individuals we had previously identified as having *NF1* PVs (7.1%) were found to harbor an additional PV in a different cancer predisposition gene (**Table S11**); these individuals were excluded from the following analyses and are discussed in **Supplemental Results**. This rate is higher than the rate of 2.9% that has previously been reported for patients found to have multiple pathogenic variants on MGPT for hereditary cancer predisposition.^29^ For the following analyses, we defined a control group, the Tested-Negative group, to include all 31,598 patients who had completed MGPT at Ambry with gene panels containing the *NF1* gene, but whose genetic testing revealed no pathogenic or likely pathogenic variants in any cancer predisposition gene.

Altogether 110 individuals (72.4%) in the Clinical-NF1 group, 103 (79.8%) in the PV-Only group, and 21,659 (70.2%) in the Tested-Negative group had a personal history of cancer (**Figure 4A**). Adjusting for patient age, this difference in incidence of malignancy between the Clinical-NF1 and PV-only group was not significant; however, compared to the Tested-Negative group, both the Clinical-NF1 (p=0.004) and PV-Only groups (p=0.03) were significantly more likely to have a personal history of cancer. Individuals in the Clinical-NF1 group also were found to have a significantly greater number of primary cancers compared to the Tested-Negative group (p=6.8e-05), whereas no difference was seen in number of primary malignancies between the Tested Negative and PV-Only groups, or between the Clinical-NF1 and PV-Only groups (**Figure 4B**). We found no significant difference between the time from cancer diagnosis to time of genetic testing between any of the three groups (**Figure S3A**). All of these trends also held true when dividing the Ambry cohort not by NF1 diagnosis status, but by *NF1* PV zygosity (*i.e.* heterozygous vs mosaic, **Figure S4A-B, S3B**).

**Figure 4:**
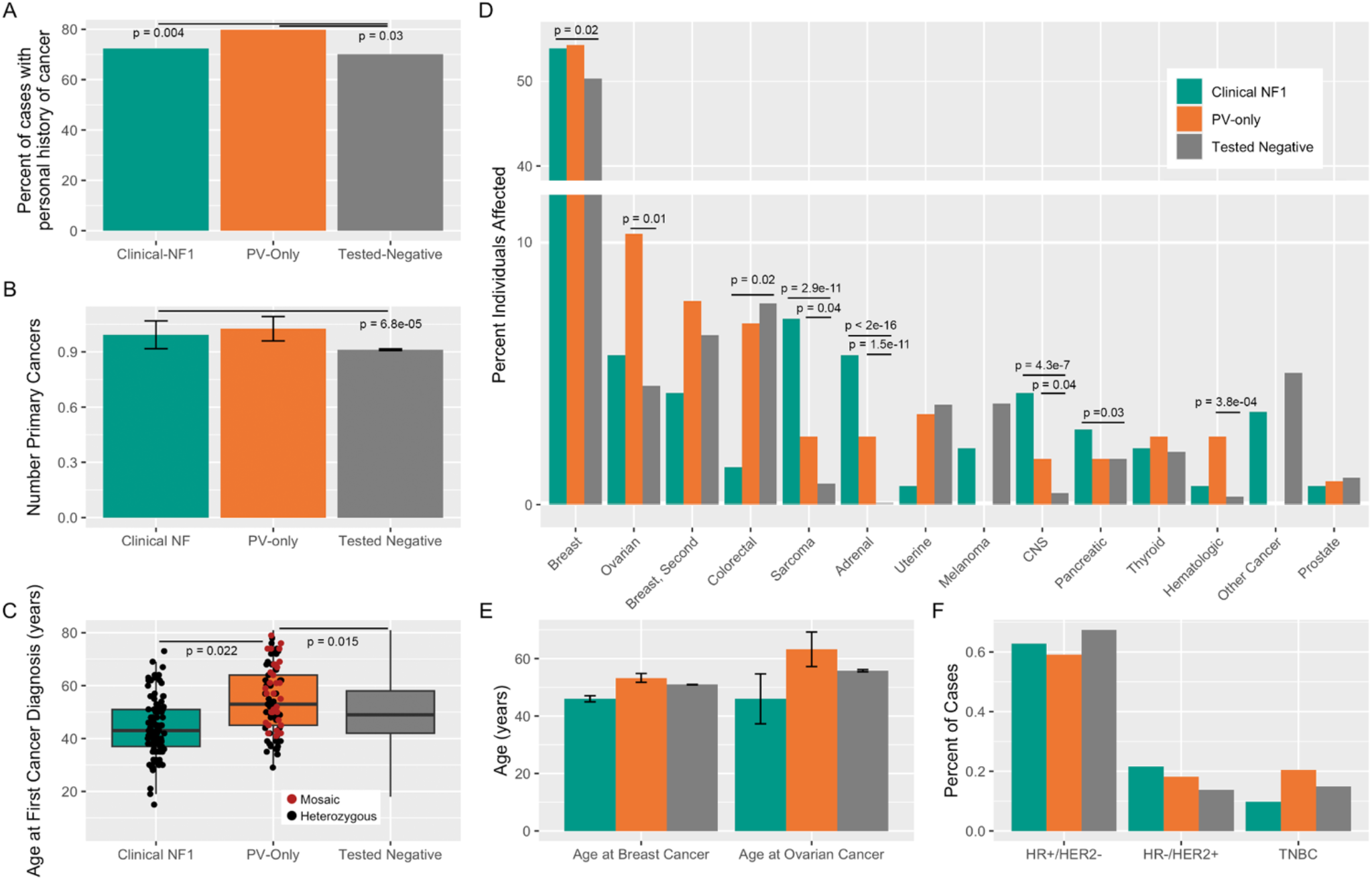
Comparison of cancer-related phenotypes between the Clinical-NF1, PV-Only, and Tested-Negative groups in the Ambry cohort. For all panels, nominally statistically significant differences between groups are labelled. In all cases, linear (for continuous response variables) or logistic regression (for categorical response variables) models were adjusted for patient age. (A) Percent of patients within each group (Clinical-NF1 in green, PV-Only in orange, and Tested-Negative in grey) reporting a personal history of malignancy. (B) Mean number of primary malignancies, per patient, across the three groups. Error bars represent standard deviation. (C) Comparison of age at first cancer diagnosis between each group. Box plots illustrate the median, first and third quartiles, minimum, and maximum for each group. For the Clincal-NF1 and PV-Only group, individual data points are shown. PV-Only individuals with mosaic *NF1* PVs are shown in red. (D) Percent of individuals, per group, affected by each of 14 different malignancies. (E) Mean age at first breast and ovarian cancer diagnosis across the three groups. Error bars represent standard deviation. (F) Incidence, across the three groups, of different breast cancer receptor statuses among patients for whom a diagnosis of breast cancer was reported and sufficient receptor status information was provided. HR, hormone receptor; HER2, human epidermal growth factor receptor 2; TNBC, triple negative breast cancer.

### Older age at cancer diagnosis for patients in the PV-Only group

Individuals in the PV-Only group were significantly older (mean 54.2 years) than both the Clinical-NF1 (mean 44.0 years) and Tested-Negative groups (mean 49.8 years) at the time of first cancer diagnosis (p=0.022 and 0.015 respectively, **Figure 4C**). Dividing the PV-Only group into two subgroups by *NF1* PV zygosity (*i.e.* heterozygous vs. mosaic), individuals with a mosaic *NF1* variant trend towards having an older age at first cancer diagnosis compared to individuals with a heterozygous variant (**Figure 4C, S4C**).

### Increased prevalence of malignancies in both the Clinical-NF1 and PV-Only groups

Dividing cancer diagnoses by type and adjusting for patient age, significant differences were seen between the three groups (**Figure 4D**, **Table S12**). Compared to the Tested-Negative group, the Clinical-NF1 group was significantly more likely to be affected by breast cancer (p=0.02), sarcoma (p=2.9e-11), adrenal cancer (p<2e-16), central nervous system (CNS) cancers (p=4.3e-07), and pancreatic cancer (p=0.03), and were significantly less likely to be affected by colorectal cancer (p=0.02) (**Figure 4D**). Patients in the PV-Only group were significantly more likely to be affected by ovarian cancer (p=0.01), sarcoma (p=0.04), adrenal cancers (p=1.5e-11), CNS cancers (p=0.04), and hematologic malignancies (p =3.8e-04) compared to the Tested-Negative group (**Figure 4D**). The increased risk for ovarian and hematologic malignancies, the rates of which were more than double what was observed in the Tested-Negative group, was unique to the PV-Only group, and was not seen in the Clinical-NF1 group. No significant differences were seen in rates of specific malignancies between the Clinical-NF1 and PV-Only groups. Again, most of these trends held true when dividing the Ambry cohort by *NF1* variant zygosity rather than NF1 diagnosis status (**Figure S4D**).

### Receptor status and age at diagnosis for breast and ovarian cancer

Individuals in the Clinical-NF1 group were diagnosed with breast cancer at younger ages (mean 46.0 years) compared to both the PV-Only (mean 53.3 years) and Tested-Negative groups (mean 51.0 years), but when adjusting for patient age at time of testing, this difference was not statistically significant (**Figure 4E**). Similarly, no statistically significant differences were seen in age at ovarian cancer diagnosis between any of the three groups (**Figure 4E**). When comparing breast cancer receptor status between the three groups, no significant differences were seen (**Figure 4F**, **Table S13**). These trends all held generally true when dividing the Ambry cohort by *NF1* variant zygosity rather than NF1 diagnosis status (**Figure S4E-F**), with the additional finding that patients with mosaic *NF1* PVs were significantly more likely to be affected by HER2+ breast cancers, compared to the Tested-Negative group (p=0.01).

## Discussion

NF1 is a classic Mendelian disorder with a characteristic phenotypic presentation.^4^ However, our understanding of the disorder is entirely based on patients ascertained through phenotype-first approaches.^30–32^ Our study, leveraging a genotype-first approach in two unique large patient cohorts, revealed surprising results: *NF1* PVs are significantly more common than would be expected given the prevalence of NF1; half of all patients with *NF1* PVs are apparently unidentified; 15-30% of all *NF1* PVs appear to be present in the somatic mosaic state; and incidentally discovered *NF1* PVs are significantly associated with an increased incidence of malignancy.

The incidental discovery of pathogenic variants is a unique challenge brought on by the advent of genomic medicine and the broad application of genetic testing in larger patient populations. Although incidentally discovered medically-actionable variants are detected in 3-6% of individuals undergoing broad genetic sequencing,^33–35^ some studies suggest that as few as 18% of individuals with incidentally discovered medically-actionable variants will have any medical history related to these genetic findings.^36^ Our results are in line with these observations, suggesting that *NF1* PVs are substantially more common than previously thought but are often discovered in apparently unaffected individuals.

Our data suggest that somatic mosaicism (both post-zygotic and clonal hematopoiesis), incomplete penetrance, and missed diagnoses all contribute to the high prevalence of *NF1* PVs that we observed. Reports on the prevalence and penetrance of Mendelian disorders such as NF1 all suffer from ascertainment bias;^37^ patients with less obvious presentations go undiagnosed, artificially decreasing estimates of disease/genetic variant prevalence. However, missed diagnoses in mildly affected patients, although likely true of some patients in our study, cannot completely explain our findings. Our experience with Cases 1-4 suggests that reduced penetrance of the disorder might be possible; *NF1* PVs can be identified in an apparent heterozygous state in the blood of patients lacking any manifestation of syndromic NF1. However, we cannot rule out the possibility that other tissues in these patients might have lower VAF of the *NF1* PV without extensive testing of different tissues. The true population prevalence of such cases is difficult to estimate from our data, which is one limitation of our study.

However, the largest single contributor to the high prevalence of *NF1* PVs that we observed was somatic mosaicism. Fifteen to thirty percent of *NF1* PV carriers in our study had low VAF *NF1* PVs with strong evidence for mosaicism in blood. This includes one individual, individual P41, who met clinical diagnostic criteria for NF1 on exam with all areas of skin affected, but whose *NF1* variant was present at only 0.10 VAF in blood, with previous clinical sequencing of the *NF1* gene being reported as negative; this case clearly illustrates the challenges posed by somatic mosaicism in clinical diagnostic workup. As we are limited in our ability to confidently call mosaic variants in all sequenced individuals due to read depth limitations and having access to only a single tissue per patient, the prevalence of mosaicism that we encountered is likely an underestimate.

Somatic mosaic *NF1* variants in the blood have previously been reported in the context of hematologic malignancy,^38–40^ and rarely in the context of clonal hematopoiesis (CH).^12, 14^ The patients that we identified with mosaic *NF1* PVs were not known to have active hematologic malignancies, arguing against this etiology as a major driver of the mosaicism that we observed. While post-zygotic mosaicism and CH are indistinguishable in the assay(s) used here, clinical context provides multiple lines of evidence argue against an age-related process like CH as the exclusive underlying etiology for the mosaicism we encountered. This finding is in line with our experience with individual P41 described above, and with a recent report of widespread mosaic genetic variants found in blood that are not clearly attributable to malignancy or CH, but likely due to post-zygotic mosaicism.^41, 42^ Thus, our data suggest that much of the apparent mosaicism that we observe for *NF1* PVs may represent post-zygotic mosaicism rather than CH, and that mosaicism for *NF1* PVs is common.

Most importantly, our results suggest that all *NF1* PVs, incidentally discovered or otherwise, are associated with increased incidence of malignancy; in the Ambry cohort, both the Clinical-NF1 and PV-only groups were significantly more likely than the Tested-Negative group to have a personal history of malignancy, even when adjusting for patient age. Although our PheWAS analysis in PMBB PV-Only individuals did not specifically replicate this finding, the overall rate of malignancy in the PMBB PV-Only group was 45.7%, higher than the reported lifetime cancer risk of 39.7% in the United States^43^ and higher than the malignancy rate of 39.1% observed in the Clinical-NF1 group (**Table S5**), a population that is known to be at increased risk of malignancy. The PMBB PV-only individuals who did have a cancer diagnosis in their charts had a wide range of malignancy types, many of which are known be associated with somatic PVs in *NF1*, further supporting the link to cancer predisposition in this group.^11^ These findings, along with our observation that both PV-Only groups are enriched for mosaic *NF1* PVs and are significantly older at the time of first cancer diagnosis, suggests that mosaic and incidental *NF1* PVs confer a real but perhaps attenuated cancer predisposition compared to germline *NF1* PVs in patients with syndromic NF1. These findings provide some guidance and prognostication for patients with incidentally discovered *NF1* variants.

It is important to note that our cohort, representing the largest known cancer-focused cohort of patients with *NF1* pathogenic variants, confirms some previous observations about associations between NF1 and malignancy and contradicts others. We confirm the previously reported younger age at first cancer diagnosis among those with heterozygous *NF1* pathogenic variants and increased incidence of breast cancer among patients with Clinical NF1 but failed to identify an earlier age of diagnosis for breast cancer or an enrichment of HER2-amplified or triple negative breast cancers in patients with NF1.^8–10^ These inconsistencies with previous work may be explained by sampling bias. The patient population that we studied, referred for genetic testing at Ambry, is likely enriched for individuals with earlier and more aggressive breast cancers (as suggested by the average age at breast cancer diagnosis of 51 years in the Tested-Negative group), and thus may not completely reflect the epidemiology of breast cancer in the general population. However, our data do corroborate the increased incidence of sarcoma, adrenal cancers, CNS malignancies, and pancreatic cancers in patients with clinical NF1; increased incidence of these same malignancies was observed in the PV-Only group, suggesting similar mechanisms of oncogenesis and predispositions to malignancy in patients even with incidentally-discovered *NF1* PVs. Intriguingly, we also observed an increased incidence of hematologic and ovarian cancers specifically within the PV-only group. Our data do not allow us to dissect the causal relationship between these observations. It is possible that incidentally discovered *NF1* PVs in the blood confer increased risk for ovarian and hematologic malignancy, as both tumor types are known to often harbor somatic mutations in *NF1*, but it also possible that these variants developed secondarily due to treatment-related CH, reflecting the relatively high rates of chemotherapy employed early in the treatment course of these malignancies.^44^

There are many limitations to our study. Our dependence on the EHR and test requisition forms for patient phenotyping is subject to error. Even with optimized data abstraction efforts the distinction between Clinical-NF1 and PV-only may be challenging. Similarly, our classification of *NF1* PVs as mosaic or germline was based on bulk sequencing of only a single tissue, and we are thus limited in our ability to confidently call mosaic variants or distinguish between post-zygotic mosaicism and CH. Although our study is the largest to investigate the incidence of malignancy in patients with *NF1* PVs, these variants are still relatively rare, and we are likely underpowered to detect small differences in patient phenotypes. Lastly, the two populations that we study in this report represent relatively sick patient populations. The Ambry cohort is enriched for patients with a personal and/or family history of malignancy. The PMBB cohort, while more reflective of the general population, was drawn from the population of patients of Penn Medicine, and is thus enriched for patients with disease. Thus, more study is required to validate the generalizability of our findings to other patient populations.

Despite these limitations, our work in two distinct and complementary patient cohorts suggests that *NF1* PVs are substantially more common than previously appreciated, are often characterized by somatic mosaicism and reduced penetrance, and are associated with increased incidence of malignancy even in patients without syndromic NF1. Although our work begins to suggest a framework for the clinical interpretation of incidentally identified pathogenic variants in the *NF1* gene, the establishment of optimal screening and management strategies will require further research and clinical efforts. It is inevitable that the continued expansion of broad genetic sequencing in large patient populations will identify incidental variants in many other genes, some of which will also be found to have important clinical associations. A broader understanding of the true population-level frequency and pathogenicity of germline and somatic mosaic variants in Mendelian disease genes across the genome will likely lead to a transformation in our understanding of the genetic architecture of both rare and common human genetic disease.

## Methods

### PMBB patient recruitment and exome sequencing

The Penn Medicine BioBank (PMBB)^25^ is a University of Pennsylvania academic biobank, approved under the University of Pennsylvania Institutional Review Board (IRB) protocol# 813913, which recruits patient-participants from the University of Pennsylvania Health System around the greater Philadelphia area in the United States. Appropriate consent was obtained from each participant regarding storage of biological specimens, genetic sequencing, access to all available EHR data, and permission to recontact for future studies. The study was approved by the Institutional Review Board of the University of Pennsylvania and complied with the principles set out in the Declaration of Helsinki. This study included the subset of 43,731 individuals enrolled in PMBB who had previously undergone exome sequencing and genotyping array. Briefly, for each individual, DNA was extracted from stored buffy coats and exome sequences were generated by the Regeneron Genetics Center (Tarrytown, NY) and mapped to GRCh38 as previously described.^45^ Sample-level filtering was as follows: individuals with low exome sequencing coverage (less than 75% of targeted bases achieving 20× coverage) or with high missingness (greater than 5% of targeted bases) were removed from analysis, leaving 43,612 samples after sample-level filtering. Variant-level filtering was as follows: in each sample, all single nucleotide variants (SNVs) with a total read depth < 7 were changed to “no-call”, and similarly all insertion/deletion (INDEL) variants with a total read depth < 10 were changed to “no-call.” Relatedness across all samples was calculated in PLINK^46^ using a minimum PI_HAT cutoff of 0.09375 to capture out to 3rd degree relationships. None of the *NF1* PV carriers that we identified were 3^rd^ degree relatives or closer.

### PMBB variant annotation

The genetic variants in PMBB exome sequencing data were subsetted to include only the *NF1* gene locus and were subsequently annotated using the Ensembl Variant Effect Predictor (VEP, version 102)^47^ with the plugin LOFTEE (version 0.3)^48^ to specifically annotate predicted loss of function (pLOF variants) and dbNSFP (version 4.2)^49^ to specifically annotate missense variants within the *NF1* gene. Only variants affecting the NCBI RefSeq^50^ canonical *NF1* transcript (NM_001042492.2) were considered. pLOF variants were defined to include nonsense variants, frameshift insertions/deletions, disruption of canonical splice site dinucleotides, or gain/loss of the start or stop codon that were not predicted to escape nonsense-mediated decay. Pathogenic missense variants were defined to include only those missense variants affecting NM_001042492.2 which had been unambiguously annotated as “pathogenic” or “likely pathogenic” in the ClinVar database. Copy number variants (CNVs) were annotated, starting with the same exome sequencing data as described above, using version 1.3 of the CLAMMS pipeline.^51^ Standard quality control measures were taken both at the sample level (samples with >40 CNVs or with >40,000 exons called as CNVs were removed) and chromosome level (for samples with >10% of a chromosome covered by >1 CNV call, that chromosome was removed). QC levels ranging from 0-3 were assigned to each CNV call based on Q_non_dip, Q_exact, and allele balance and heterozygosity metrics. Only CNVs meeting the most stringent QC threshold of 3 were included in the analysis. The resultant CNV calls at the *NF1* locus were manually reviewed to identify any potentially suspicious annotations – no CNVs were excluded after manual review. The data available in PMBB did not permit us to determine VAF or potential mosaicism for CNVs.

### PMBB chart review

Chart review was carried out under the University of Pennsylvania Institutional Review Board (IRB) protocol 853693 and was found to meet criteria for IRB review exemption. We performed manual chart review of complete EHR data for each carrier of an *NF1* pathogenic variant that we identified in PMBB. All 58 charts were reviewed by a single clinician blinded to the results of exome sequencing, and detailed information was extracted. At a minimum for each individual we gathered: any mention of the terms “NF1” “neurofibromatosis” or “von Recklinghausen;” self-reported race; height at most recent measurement; number of encounters with UPHS providers (if fewer than 5); whether the individual had been seen by a clinical geneticist; all documented skin exams; all documentation regarding malignancies. The clinician subsequently generated a summary paragraph describing each individual’s overall health issues and diagnoses. This information is documented in **Table S2** and was used to generate the summary statistics listed in **Table S3**.

### PMBB phenotype generation

ICD-9 and ICD-10 disease diagnosis codes and procedural billing codes were extracted from patient EHRs cross the entire PMBB dataset. ICD-9 encounter diagnoses were mapped to ICD-10 using the Center for Medicare and Medicaid Services 2017 General Equivalency Mappings (https://www.cms.gov/Medicare/Coding/ICD10/2017-ICD-10-CM-and-GEMs.html) with unmappable ICD-9 codes dropped from further analysis. Participants were determined as having a phenotype if they had the corresponding ICD diagnosis code on at least one date, while phenotypic controls consisted of individuals who never had the ICD code used. ICD codes S00-T98 (within the group “Injury, poisoning and certain other consequences of external causes”) and V01-Y98 (within the group “External causes of morbidity and mortality”) were excluded from PheWAS analysis. Only ICD phenotypes with a total case count of 20 or more across all 43,612 individuals in the analysis were included in our study.

### PMBB Phenome-wide association studies (PheWAS)

Using a PheWAS^52^ approach, all *NF1* pathogenic variants in PMBB were collapsed into a single binned genotype and tested for association with each ICD code-derived phenotype (described above) using the program BioBin^53^ with a logistic regression model adjusted for age, sex and the first ten principal components of genetic ancestry. Our association analyses considered only phenotypes with at least 15 cases across all of PMBB, as described above, leading to the interrogation of 9,032 total phenotypes. Subsequent PheWAS analyses were completed using the same approach, but excluding from the analysis either the patients in the Clinical-NF1 group or PV-Only group. Q-Q plots were generated to assess for p-value inflation (**Figure S5**). Note – 24 individuals in PMBB were identified as having ICD code Q85.01 (Neurofibroatosis, type 1), but only 18 of these were found to have an NF1 PV in our analysis. The remaining 6 individuals with ICD code Q85.01 may have had smaller CNVs not detectable by our methodology, missense variants not classified as pathogenic/likely pathogenic, deep intronic variants missed on exome sequencing, or even ICD codes entered in error.

### Ambry cohort definition and molecular testing

We queried a cohort of 118,768 patients from the Ambry Genetics (Aliso Viejo, CA) laboratory database from 1/1/2014 through 3/31/2018 who had undergone clinical Next-Generation Sequencing (NGS) to identify individuals with pathogenic or likely pathogenic variants in *NF1*. This cohort contained sequencing data obtained from clinical multigene panel testing, including BreastNext, CancerNext, CancerNext Expanded, OvaNext, CustomNext, PGLNext, and BrainTumorNext, covering between two and 67 genes related to hereditary cancer risk (**Table S4).** All patients were clinician referred; ordering standards were based on clinician judgement or practice-specific thresholds. Sequencing, variant calling, and variant annotation was performed at Ambry Genetics as previously described^29^ and all identified pathogenic/likely pathogenic *NF1* variants were classified as described in **Table S5**. Pathogenic and likely pathogenic variants (PVs) were defined to include deletions (including whole gene deletions or smaller deletions encompassing at least one exon of the *NF1* gene), exonic duplications, frameshift variants, nonsense variants, canonical splice site variants, and missense variants meeting ACMG/AMP criteria for pathogenicity.^54^ Regions with <20× coverage on NGS were followed up with Sanger analysis. In addition, variants in regions complicated by pseudogene interference, variant calls not satisfying depth of coverage (100x) and variant allele frequency (40%) quality thresholds, reportable small insertions and deletions, and potentially homozygous variants were verified by Sanger sequencing. Standard protocols for clinical testing were used for automated variant calling in Sanger sequencing (SeqPilot, JSI Medical Systems) analysis. Where variants were not called by the software but clearly visible at low levels on visual inspection by qualified personnel, variants were designated as mosaic

### Ambry cohort patient phenotyping

For the Ambry data set, demographic and clinical information including gender, personal cancer history, family cancer history, and family history of Neurofibromatosis type 1 (NF1) was obtained through review of laboratory test requisition forms (TRF) and, where available, clinic notes and pedigrees. For cases lacking documentation of a clinical diagnosis of NF1, referring providers were contacted to obtain further clinical details. Patients were subsequently categorized as “Clinical-NF1” or “PV-only” (pathogenic variant-only) using a three-tiered classification process (**Figure S1, Supplemental Results**). First, when physical exam data was documented, cases of Clinical-NF1 were defined using the NIH NF1 diagnostic criteria.^55^ Second, if physical examination data were not recorded but the patient had a documented first-degree family member with NF1, the patient was included in the Clinical-NF1 group. Third, if neither physical exam nor family history data were available, patients were categorized according to clinician-provided description on the TRF. PV-Only cases were ideally defined by comprehensive physical exam data documenting lack of concordance with NIH criteria (as illustrated in the top branch of **Figure S1**). For most cases, however, PV-Only cases were defined by information in the TRF.

### Statistical analysis

All statistical analyses were performed with R software, version 4.2.2. Within both the PMBB and Ambry cohorts, differences in *NF1* PV VAF and patient age between the Clinical-NF1 and PV-Only groups were compared by linear regression. Differences in the distributions of individual ages between the *NF1* PV-Only group and the overall PMBB/Ambry datasets were compared using the Wilcoxon rank sum test. Note that for the 2 whole-gene deletions identified in the PMBB cohort, and for 25 individuals in the Ambry cohort, VAF could not be determined, and these individuals were excluded from analyses requiring VAF. For all categorical traits and phenotypes listed in **Table 5** and **Table S6**, statistical analyses were completed by logistic regression (logistic regression models were adjusted for patient age and sex as indicated). For the continuous traits of VAF, read depth, height, and age, statistical analyses were accomplished via linear regression. The proportions of each class of genetic alteration and mutational spectrum were compared between groups with logistic regression. Within the Ambry cohort, comparisons between the Clinical-NF1, PV-Only, and Tested-Negative groups, or between the heterozygous *NF1* PV, mosaic *NF1* PV, and Tested-Negative groups, were completed as follows. Age at first cancer diagnosis, number of cancer primaries, age at first breast cancer diagnosis, and first ovarian cancer diagnosis were compared between the groups by linear regression, adjusting models for individual age at the time of testing. The incidence of personal history of cancer, of each specific malignancy, and breast cancer receptor status, were compared by logistic regression adjusting the model for individual age at the time of testing. Time between first cancer diagnosis and genetic was compared using the Wilcoxon rank sum test.

## Data Availability

All data produced in the present work are contained in the manuscript

## Acknowledgments

We would like to thank members of the Ritchie Lab who provided scripts or analysis support for aspects of these analyses. We acknowledge the Penn Medicine BioBank (PMBB) for providing data and thank the patient-participants of Penn Medicine who consented to participate in this research program. We would also like to thank the Penn Medicine BioBank team and Regeneron Genetics Center for providing genetic variant data for analysis. The PMBB is approved under IRB protocol# 813913 and supported by Perelman School of Medicine at University of Pennsylvania, a gift from the Smilow family, and the National Center for Advancing Translational Sciences of the National Institutes of Health under CTSA award number UL1TR001878. T.G.D. is supported in part by the NIH K08 1K08DK127247 and by the Burroughs Wellcome Fund. M.D.R. is supported in part by NIH R01 AI077505, GM115318, AI116794. KLN is supported in part by the Breast Cancer Research Foundation, Basser Center for BRCA, and the National Institutes of Health -R01 CA176785 and R01 CA192393.

## SUPPLEMENTAL INFORMATION

### Supplemental Results

#### Classification of patients into Clinical-NF1 and PV-Only groups in the Ambry cohort

We divided the Ambry cohort into two groups – 1) those having an *NF1* PV and a known diagnosis of NF1 (the Clinical-NF1 group), and 2) those with an *NF1* PV but without evidence of having NF1 (the PV-only group). The approach used to make this distinction was as follows (**Figure S1**). Thirty-seven of the 275 *NF1* PV carriers (14%) had sufficient physical exam documentation in their available clinic notes to inform our classification; 34 of these met NIH diagnostic criteria for NF1, while three had sufficiently documented physical exam data to demonstrate that they did not meet NIH criteria. Thirty of the 34 patients (88%) with well-documented physical exam findings leading to their classification as having Clinical-NF1 also had NF1 mentioned on the test requisition forms (TRF), demonstrating high concordance between TRF and physical exam documentation. An additional 16 *NF1* PV carriers (6%) did not meet NIH diagnostic criteria based on TRF/clinic note review but were classified as clinical-NF1 due to a documented family history of NF1. Lastly, an additional 102 *NF1* PV carriers (37%) were classified as clinical-NF1 based on mention of NF1 in the TRF. Thus, altogether, 152 patients (54%) were classified as having clinical-NF1, whereas 129 patients (46%) were classified as PV-Only.

#### Patients in the Ambry cohort with concurrent pathogenic variants in *NF1* and additional cancer predisposition genes do not have an earlier age of cancer diagnosis

Of the 152 cases in the Clinical-NF1 group, eight (5.3%) had concurrent pathogenic variants identified in additional cancer predisposition genes; of the 129 patients in the PV-Only group, concurrent pathogenic variants were found in 12 (9.3%). This difference was not significant by chi-square analysis (p=0.19). The genes involved, as well as the patients’ personal oncologic history, family history, and concurrence with NCCN guidelines for genetic testing are detailed in **Table S11**. We did not find a difference between the mean age at cancer diagnosis in patients with an *NF1* pathogenic variant alone and patients with additional concurrent pathogenic variants in other cancer predisposition genes; for patients with Clinical-NF1 with pathogenic variants in the *NF1* gene only, the mean age at cancer diagnosis was 43.9, while the mean age at cancer diagnosis was 42.4 years for patients with additional cancer susceptibility gene pathogenic variants. For patients in the PV-Only group, these ages were 53.9 years and 54.6 years, respectively. We excluded all 20 patients with pathogenic variants in genes outside of *NF1* from subsequent malignancy-related analyses.

## PENN MEDICINE BIOBANK BANNER AUTHOR LIST AND CONTRIBUTION STATEMENTS

### PMBB Leadership Team

Daniel J. Rader, M.D., Marylyn D. Ritchie, Ph.D.

Contribution: All authors contributed to securing funding, study design and oversight. All authors reviewed the final version of the manuscript.

### Patient Recruitment and Regulatory Oversight

JoEllen Weaver, Nawar Naseer, Ph.D., M.P.H., Afiya Poindexter, Khadijah Hu-Sain, Yi-An Ko, Ph.D.

Contributions: JW manages patient recruitment and regulatory oversight of study. NN manages participant engagement, assists with regulatory oversight, and researcher access. AP, KH, YK perform recruitment and enrollment of study participants.

### Lab Operations

JoEllen Weaver, Meghan Livingstone, Fred Vadivieso, Stephanie DerOhannessian, Teo Tran, Julia Stephanowski, Monica Zielinski, Ned Haubein, Joseph Dunn

Contribution: JW, ML, FV, SD conduct oversight of lab operations. ML, FV, AK, SD, TT, JS, MZ perform sample processing. NH, JD are responsible for sample tracking and the laboratory information management system.

### Clinical Informatics

Anurag Verma, Ph.D., Colleen Morse Kripke, M.S. DPT, MSA, Marjorie Risman, M.S., Renae Judy, B.S.

Contribution: All authors contributed to the development and validation of clinical phenotypes used to identify study subjects and (when applicable) controls.

### Genome Informatics

Anurag Verma Ph.D., Shefali S. Verma, Ph.D., Yuki Bradford, M.S., Scott Dudek, M.S., Theodore G. Drivas, M.D., Ph.D.

Contribution: A.V., S.S.V. are responsible for the analysis, design, and infrastructure needed to quality control genotype and exome data. Y.B. performs the analysis. T.G.D. and A.V. provides variant and gene annotations and their functional interpretation of variants.

## Supplemental Figures

**Figure S1:**
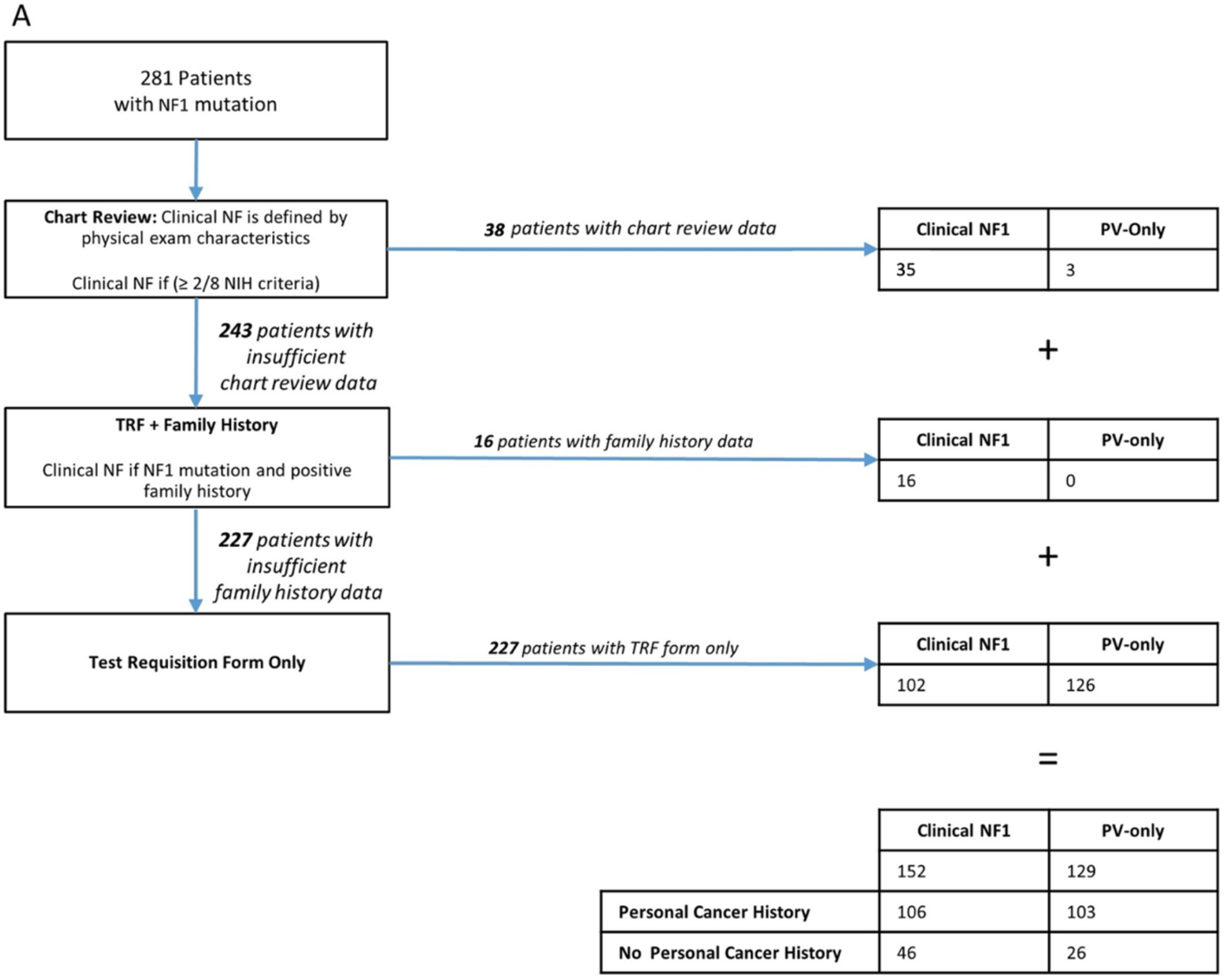
Flow diagram of patients with NF1 pathogenic and likely pathogenic variants outlining how clinical diagnosis was determined. Of the 281 patients found to have pathogenic variants in *NF1* in the Ambry data set, one hundred and twenty-nine patients (46%) were classified as “PV-only”, whereas 152 patients (54%) were classified as having “clinical NF1”. A total of 38 (35 clinical NF1 and 3 PV-only) cases between these two groups were made on the basis of documented physical exam concordance with NIH criteria. Furthermore, 15 patients did not meet NIH criteria, but were classified as “clinical NF1” due to documented family history. The remainder of the patients (228) were therefore classified based on test requisition form (TRF) alone.

**Figure S2:**
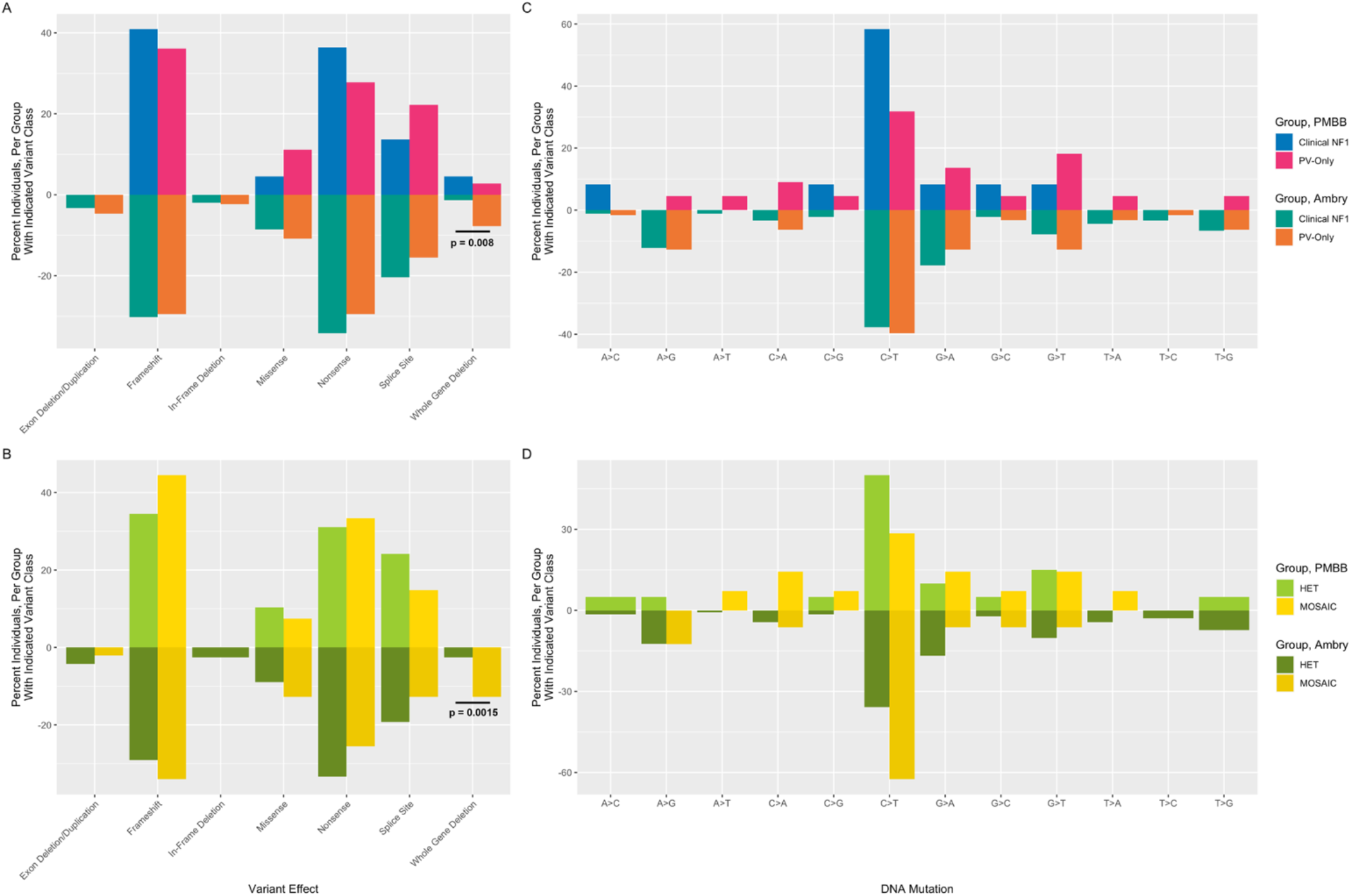
Comparison of different *NF1* variant types between the PMBB and Ambry Clinical-NF1 and PV-Only groups. The percent of individuals in each dataset (PMBB in the top half of each graph, Ambry in the bottom half) harboring different classes of *NF1* variants, are displayed. In all cases, the PMBB mosaic *NF1* PV group was defined to include all individuals with an *NF1* PV with VAF < 0.3. The Ambry mosaic group was defined as discussed in the main text. All statistically significant differences, as determined by logistic regression, are indicated on the graph. (A) Comparison of different predicted protein effects of each *NF1* PV between the Clinical-NF1 and PV-Only groups. The PV-Only group is significantly enriched for whole gene deletions in the Ambry cohort. (B) Comparison of different predicted protein effects of each *NF1* PV between the heterozygous (HET) and mosaic (MOSAIC) *NF1* PV groups. The mosaic group is significantly enriched for whole gene deletions in the Ambry cohort. (C) For individuals with single nucleotide substitutions, the percent of individuals in each dataset harboring each specific substitution is displayed for the Clinical-NF1 and PV-Only groups. (D) For individuals with single nucleotide substitutions, the percent of individuals in each dataset harboring each specific substitution is displayed for the heterozygous and mosaic *NF1* PV groups.

**Figure S3:**
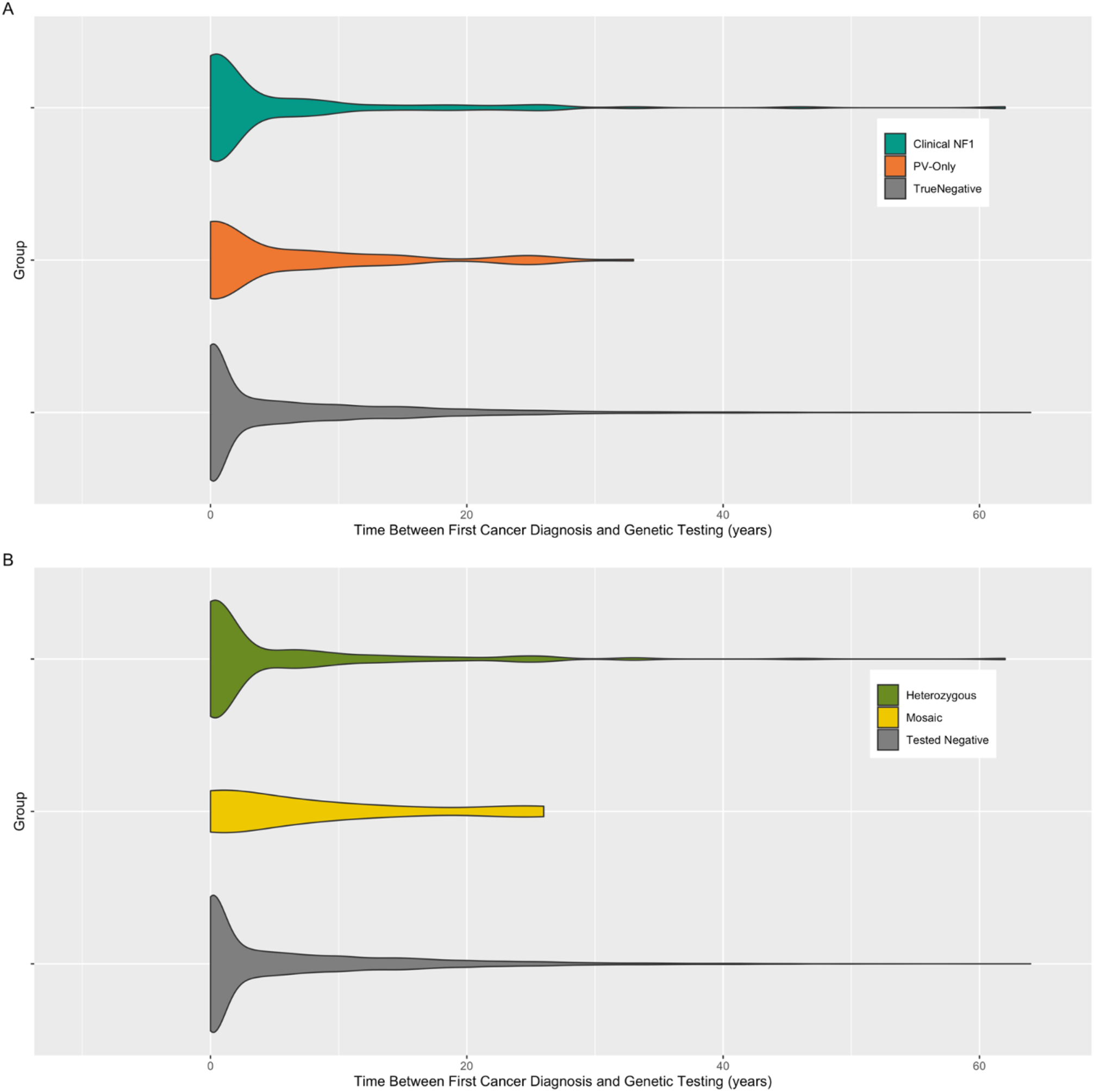
Time between first cancer diagnosis and genetic testing in the Ambry dataset. The time between an individual’s first reported cancer diagnosis, and the time at which they underwent genetic testing at Ambry, in years, is depicted using violin plots, comparing (A) the Clinical-NF1, PV-Only, and Tested-Negative groups and (B) the heterozygous *NF1* PV, mosaic *NF1* PV, and Tested-Negative groups. No statistically significant differences were found between any groups.

**Figure S4:**
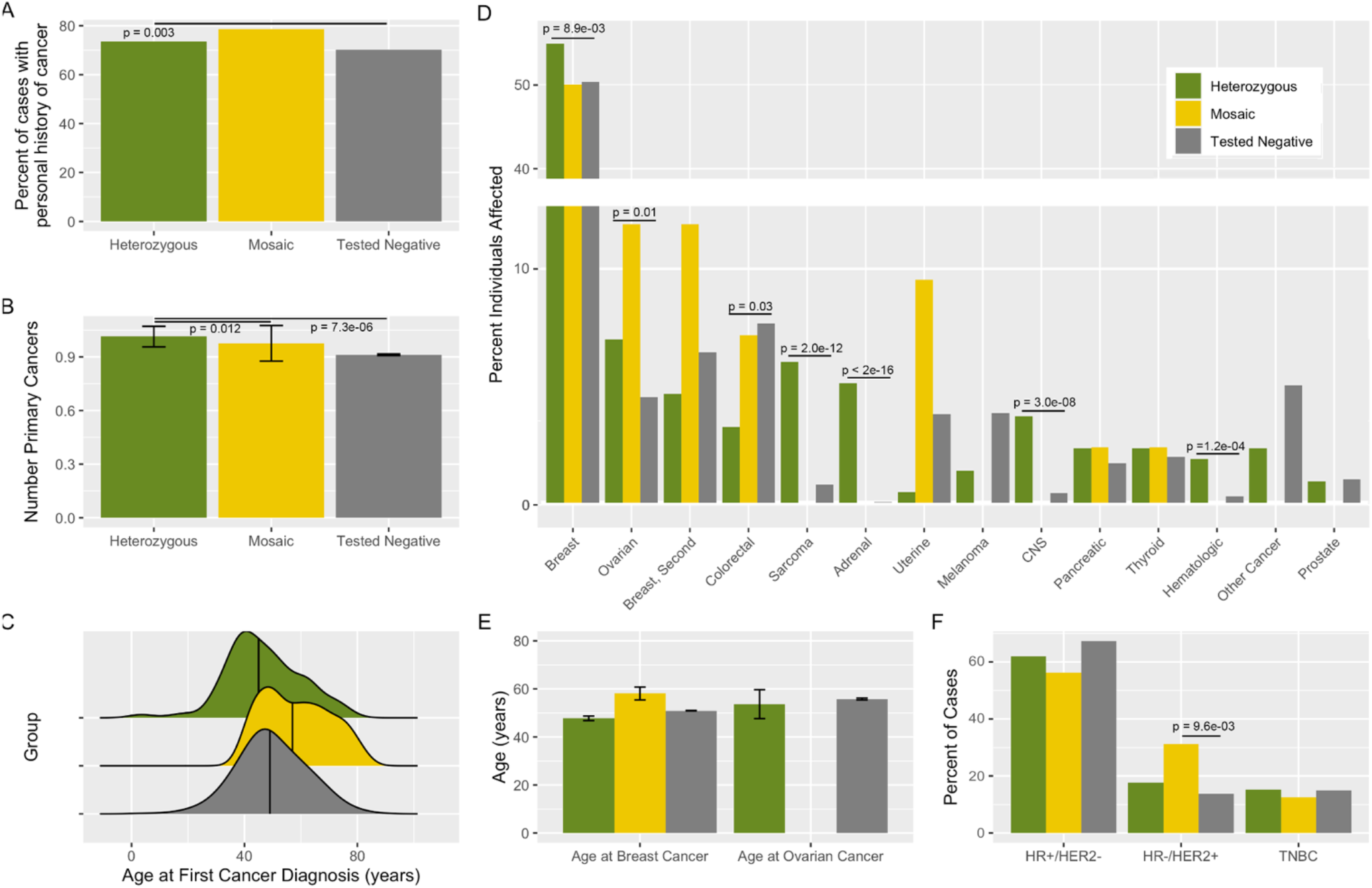
Comparison of cancer-related phenotypes between the heterozygous *NF1* PV, mosaic *NF1* PV, and Tested-Negative groups in the Ambry cohort. For all panels, nominally statistically significant differences between groups are labelled. In all cases, linear (for continuous response variables) or logistic regression (for categorical response variables) models were adjusted for patient age. (A) Percent of patients within each group (heterozygous *NF1* PV in chartreuse, mosaic *NF1* PV in yellow, and Tested-Negative in grey) reporting a personal history of malignancy. (B) Mean number of primary malignancies, per patient, across the three groups. Error bars represent standard deviation. (C) Distribution of patient age at first cancer diagnosis across the three groups. Vertical line represents the mean value, per group. (D) Percent of individuals, per group, affected by each of 14 different specific types of malignancies. (E) Mean age at first breast and ovarian cancer across the three groups. Error bars represent standard deviation. Age at ovarian cancer diagnosis was not available for any of the individuals in the mosaic *NF1* PV group (F) Incidence, across the three groups, of different breast cancer receptor statuses among patients for whom a diagnosis of breast cancer was reported and sufficient receptor status information was provided. HR, hormone receptor; HER2, human epidermal growth factor receptor 2; TNBC, triple negative breast cancer.

**Figure S5:**
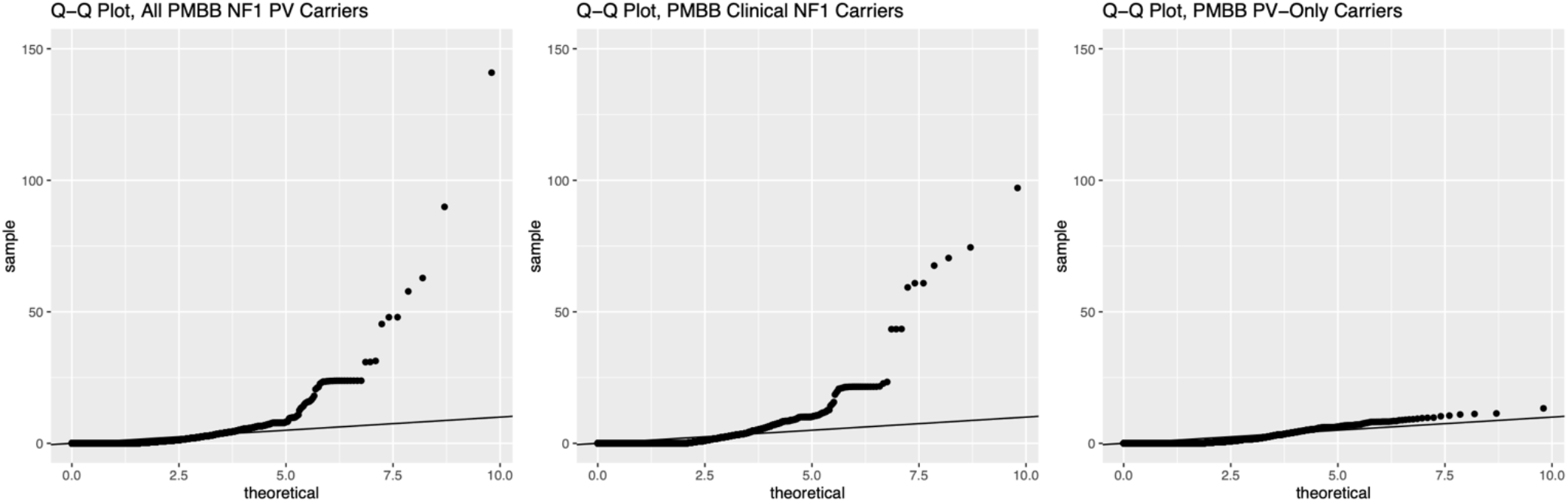
Q-Q plots for PheWAS analyses in Figure 3. Q-Q plots for the three PheWAS analyses in Figure 3 are displayed, illustrating an absence of significant artifactual p-value inflation.

## List of Supplemental Tables

Table S1. Phenotypes of patients evaluated in our clinic with incidentally discovered *NF1* pathogenic/likely pathogenic variants

Table S2. Summary of PMBB patients with *NF1* variants Table S3. Multi-gene panels included in Ambry cohort Table S4. Summary of Ambry patients with *NF1* variants

Table S5. Contingency tables and statistical analysis for different characteristics of Clinical-NF1 and PV-Only patients in PMBB

Table S6. Contingency tables and statistical analysis for different characteristics of Clinical-NF1 and PV-Only patients in Ambry cohort Table S7. Characteristics of Clinical-NF1 Patients in the Ambry Cohort with Low Variant Allele Fraction

Table S8. Summary Statistics for PheWAS Performed on all 58 *NF1* Pathogenic Variant Carriers in PMBB

Table S9. Summary statistics for PheWAS performed only on the 23 Clinical-NF1 individuals in the PMBB cohort

Table S10. Summary statistics for PheWAS performed only on the 35 PV-Only individuals in the PMBB cohort Table S11. Clinical and family history characteristics of individuals in the Ambry cohort with pathogenic variants in *NF1* and an additional cancer predisposition gene

Table S12. Counts, rates, and statistical analysis of different malignancies across Ambry patient groups

Table S13. Breast cancer receptor status in the Ambry cohort

